# Determinants and causes of early neonatal mortality: Hospital based retrospective cohort study in Somali region of Ethiopia

**DOI:** 10.1101/2022.03.16.22272337

**Authors:** Ahmed Tahir Ahmed, Abdifatah Elmi Farah, Hussein Nooh Ali, Muse Obsiye Ibrahim

## Abstract

**Background:** Early neonatal mortality occurs when a newborn dies within the first seven days of life. Despite interventions, newborn mortality in Ethiopia has grown over time (33 death per 1000 live births). Determinants varies on level of neonatal mortality. The study’s goal was to determine magnitude of early newborn death, as well as its causes and drivers, in Newborn Intensive Care Unit of Referral hospital in Ethiopia’s Somali region.

**Methods:** Health facility based retrospective study review was conducted between May 2019 to May 2021 in Shiek Hassan Yabare Referral Hospital of Jigjiga University of Ethiopia. All neonates between 0 to 7 days admitted at NICU and get registered using the new NICU registration book from May 2019 to May 2021 with complete data were included. Kobo toolkit was used for data collection and analyzed in SPSS 20. Logistic regression model was used to estimate determinants.

**Result:** The magnitude of early neonatal mortality rate (defined as death between 0-7 days) of Ethiopia’s Somali region is estimated to be 130 per 1000 live births—That is say 130 newborn couldn’t celebrate their seventh day in every 1000 live births. Hypothermia, prematurity, maternal death at birth and shorter length of stay in NICU were increasing the chance of neonatal mortality at early stage while neonatal resuscitation had shown protective effect against neonatal mortality. Similarly birth asphyxia, preterm, sepsis, and congenital abnormalities were major causes of admission and death in the NICU.

**Conclusion:** The magnitude of early neonatal mortality is considerable and causes are preventable. Enhancing quality of care including infection prevention and hypothermia through mentorship and encouraging the Kangaroo Mother Care practice is necessary at childbirth and NICU of the Hospital.

## Background

Early neonatal mortality occurs when a newborn dies within the first seven days of life [1]

Neonatal mortality continues to be the highest (17%) among children under the age of five worldwide, with Sub-Saharan Africa accounting for the largest share [2]. Low-income countries continue to have higher rates of neonatal death [3, 4].

Despite interventions, newborn mortality in Ethiopia has grown over time (33 per 1000 live births)[5]. However, a comprehensive study and meta-analysis found that early newborn mortality is reducing in Ethiopia, despite the high rate of early neonatal mortality (29.5 per 1000)[6].

Furthermore, even newborns admitted to sub-Saharan nations’ neonatal intensive care units (NICU) [7-10], including Ethiopia, have a greater rate of early neonatal mortality. The causes of newborn mortality are numerous and vary depending on the situation. Prematurity and congenital anomalies [1] and low APGAR score [11] are the leading causes of early newborn mortality in the developed world, whereas very low birth weight [9, 12] sepsis [13] and asphyxia [7, 9, 10, 12],[14] are the leading causes in the low-income world, according to evidence from systematic reviews.

In low-income countries such as Ethiopia, early newborn mortality may also be caused by insufficient access to care [7, 15, 16], home birth [16], and issues with NICU care quality [4, 8].Prematurity-related problems and late breast-feeding initiation are also causes of infant death in low-income and resource-constrained countries [7, 13, 14, 17].

Furthermore, maternal illness and death, as well as a poor Apgar score, are two additional causes of neonatal mortality [7, 12, 13, 18]. Maternal child spacing, on the other hand, protects against it [6].Finally, duration of stay is another driver in Ethiopia, particularly in pastoralist communities, in addition to the other primary causes of newborn mortality in low-income countries like Ethiopia [8, 19].

The severity of early newborn death in Ethiopia is poorly understood, and its causes and predictors are varies in different context. The study’s goal was to determine the scale of early newborn death, as well as its causes and drivers, in NICUs of health facilities in Ethiopia’s Somali region.

## Methods

### Study setting and period

Health facility based retrospective study review was conducted between May 2019 to May 2021 in Shiek Hassan Yabare Referral Hospital (SHYRH) of Jigjiga University. SHYRH began delivering services in 2017 and it aimed to give serves about 5 million people in Somali region of Ethiopia. The hospital has 400 beds and serves 86000 outpoint annually. It is the largest hospital with a number of specialties including neonatal intensive care unit (NICU). The number of average deliveries per month is 400.

### Eligibility criteria

Neonates died or survived between 0 to 7 days were reviewed among neonates admitted in NICU of SHYRH from May 2019 to May 2021. All neonates between 0 to 7 days admitted at NICU and get registered using the new NICU registration book with complete data from May 2019 to May 2021 were included. Early neonatal death was considered a newborn death within the first weeks of life (between 0 to 7 days) while in NICU[1]. Admitted neonates stayed more than 7 days in NICU and incomplete data were excluded.

### Sample size

Between April and May of 2021, an exhaustive review of the NICU registration book from May 2019 to May 2021 was conducted, with a total of 765 eligible neonates (99 deaths and 666 survivors) included in the study.

### Data collection methods

The data was collected using checklist extracted from the new NICU registration book as it is comprehensive and filled properly. Two neonatal Nurse by profession were collected the data using Kobo toolbox software to prevent errors happen during data collection. Data collectors were sending the collected data per day to the server and getting on spot feedback from the supervisors. The outcome the study was early neonatal death and determinant variables were maternal and neonatal characteristics at NICU of SHYRH.

### Data Management and Analysis

For data entry and collection, the Kobo toolkit was used. The data was cleaned with the Kobo toolkit before being analyzed with SPSS version 20. To find factors of early newborn death, researchers used bivariate and multivariable binary logistic regression. In multivariate analysis, variables having a P-value less than 0.2 in bivariate analysis, as well as additional relevant determinants of early newborn mortality, were evaluated. In multivariate binary logistic regression analysis, factors having a P-value less than 0.05 were determined to have a significant relationship with the outcome variable. Finally, the odds ratio with 95 percent confidence interval was used to estimate the strength of the association.

Furthermore, to have a better knowledge of neonatal death, a frequency distribution table was employed to discover the causes of early neonatal mortality.

### Ethical consideration

The study was ethically approved by ethical committee of college of medicine and health science of Jigjiga University, Ethiopia. Since data was collected from NICU registration book of SHYRH, an official consent was obtained from Hospital administration.

## Result

### Newborn characteristics at NICU

A total of eligible 765 neonates with complete data were reviewed and included in the study to estimate the rate of early neonatal mortality in Ethiopia’s Somali region based on neonates admitted to the Shiek Hassen Referral Hospital’s NICU. Among 765(between 0-7days) reviewed, 99 died early and 666 survived. Higher proportion 41 (41.4%) of small babies (below2000gm) died relative to 184(27.6%) survived. similarly, 88 (88.9%) and 72(72.7%) of the 99 neonates who died early were hypothermic and not resuscitated respectively. Regarding the first minute APGAR scores, mortality were higher among neonates with lower score 88(88.9%) compared to those with higher score 11(11.1%).

Survival rate was lower among neonates admitted with premature 63(9.5%), asphyxia 143(21.5%) and congenital anomalies 65(9.8%) compared to counterparts (Table 1).

**Table 1.** Characteristics of NICU admitted neonates at Referral Hospital, Jigjiga from May 2019-2021

| Newborn Characteristics |  | Discharge status |  |
| --- | --- | --- | --- |
|  |  | Death (%),<br>n=99 | Survived (%),<br>n=666 |
| Sex of the baby | Male | 54(54.5%) | 363(54.5%) |
|  | Female | 45(45.5%) | 303(45.5%) |
| Birth Weight | <2000gm(small baby) | 41(41.4%) | 184(27.6%) |
|  | 2000-2499.99gm(LBW) | 20(20.2%) | 74(11.1%) |
|  | ≥4000gm(large baby) | 1(1.0%) | 22(3.3%) |
|  | 2500-3999.99 gm(Normal) | 37(37.4%) | 386(58.0%) |
| Temperature | Normal(36.5-37.5) | 10(10.1 %) | 223(33.5%) |
|  | Low temperature(<36.5) | 88(88.9%) | 416(62.5%) |
|  | High temperature(>37.5) | 1(1.0%) | 27(4.1%) |
| APGAR Score 5<sup>th</sup><br>minute | 1-6 | 58(58.6%) | 385(57.8%) |
|  | 7-8 | 37(37.4%) | 248(37.2%) |
|  | 9 | 4(4.0%) | 33(5.0%) |
| APGAR Score 1<sup>th</sup><br>minute | 0-6 | 88(88.9%) | 547(82.1%) |
|  | 7-9 | 11(11.1%) | 119(17.9%) |
| Length of stay | ≤ 1 day | 4(4.0%) | 12(1.8%) |
|  | 2-7 days | 94(94.9%) | 498(74.8%) |
|  | 8+ days | 1(1.0%) | 156(23.4%) |
| Resuscitation | No | 72(72.7%) | 593(89.0%) |
|  | Yes | 27(27.3%) | 73(11.0%) |
| Prematurity | No | 67(67.7%) | 603(90.5%) |
|  | Yes | 32(32.3%) | 63(9.5%) |
| Sepsis | No | 55(55.6%) | 310(46.5%) |
|  | Yes | 44(44.4%) | 356(53.5%) |
| Respiratory Distress<br>Syndrome (RDS) | No | 34(34.3%) | 272(40.8%) |
|  | Yes | 65(65.7%) | 394(59.2%) |
| Perinatal Asphyxia | No | 67(67.7%) | 523(78.5%) |
|  | Yes | 32(32.3%) | 143(21.5%) |
| Congenital malformation | No | 89(89.9%) | 601(90.2%) |
|  | Yes | 10(10.1%) | 65(9.8%) |

### Maternal characteristics of neonates admitted to NICU of referral hospital

Many maternal characteristics were taken into account as determinants. Survival rate was lower among neonates born at home, by instrumental delivery and their mother died during childbirth relative to counterparts (Table 2).

**Table 2.** Maternal characteristics of neonates admitted to NICU of Shiek-Hassen Referral Hospital from May 2019-May 2021

| Maternal Characteristics |  | Discharge status |  |
| --- | --- | --- | --- |
|  |  | Death (%), n=99 | Survived (%), n=666 |
| Mode of delivery | CS | 23(23.2 %) | 182(27.3%) |
|  | Instrumental | 1(1.0%) | 22(3.3%) |
|  | Spontaneous | 75(75.8%) | 462(69.4%) |
| Place of delivery | Home delivery | 3(3.0%) | 20(3.0%) |
|  | Referred from Other | 17(17.2%) | 158(23.7%) |
|  | Same facility | 79(79.8%) | 488(73.3%) |
| HIV test | Negative | 98(12.9%) | 661(87.1%) |
|  | Positive | 1(16.7%) | 5(83.3%) |
| Hepatitis B test | Negative | 96(97.0%) | 649(97.4%) |
|  | Positive | 3(3.0%) | 17(2.6%) |
| Syphilis test | Negative | 99(100.0%) | 664(99.7%) |
|  | Positive | 0(0.0%) | 2(0.3%) |
| Maternal status | Alive | 77(77.8%) | 661(99.2%) |
|  | Dead | 22(22.2%) | 5(0.8%) |

### Magnitude of early neonatal mortality

The magnitude of early neonatal mortality rate (defined as death between 0-7 days) of Ethiopia’s Somali region is estimated to be 95% CI: 130 (106, 154) per 1000 live births. That is 130 newborn couldn’t celebrate its sevens day in every 1000 live births.

### Determinants of early neonatal mortality at NICU of Shiek Hassen Referral hospital

All newborn and maternal characteristics with a P-value less than 0.2 were considered in multivariate analysis. However; maternal status, preterm, newborn resuscitation, hyperthermia, and newborn shorter length of stay in NICU were revealed to be determinants of early neonatal mortality. When compared to those whose moms were still alive, newborns whose mothers died during childbirth were considerably more likely to die early at neonatal age (adjusted odd ratio (AOR) = 29.3; 95% CI: 9.3, 92.1). Similarly, Staying in the NICU for a short period of time (less than 24 hours) was associated with a higher rate of early neonatal mortality (AOR= 72.5; 95 % CI: (6.3, 834.2) than those who stay longer. Furthermore, compared to term born newborns and neonate with normal temperature, premature and hypothermic babies were almost four times more likely to die early at neonatal age (AOR= 3.6; 95 % CI: (1.7, 8) and AOR= 3.8; 95 % CI: (1.8, 8.1) respectively. Newborn resuscitation, on the other hand, has a protective effect and reduces early newborn death by 70% AOR=0.3; (0.2, 0.8). (Table 3).

**Table 3.**
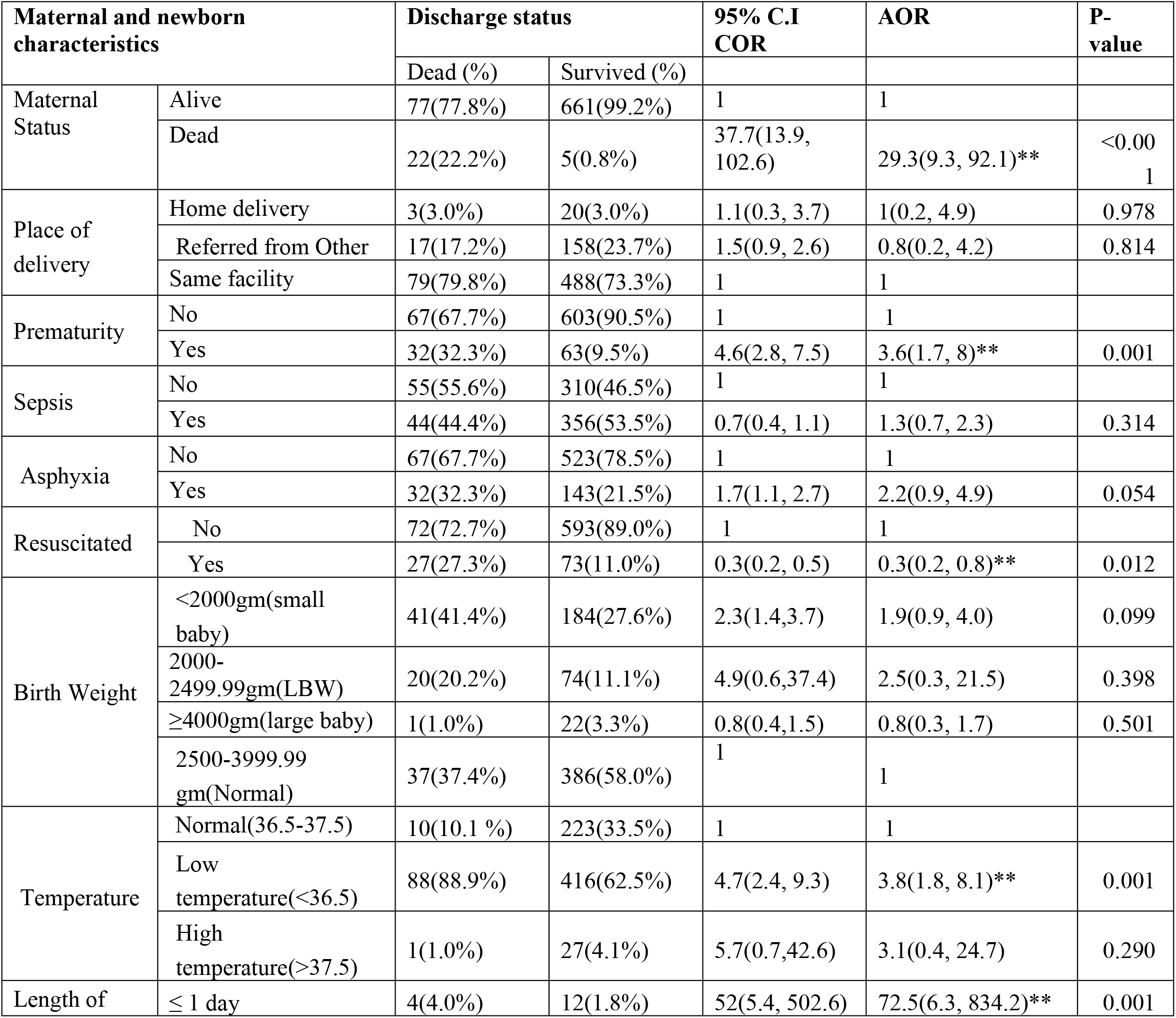
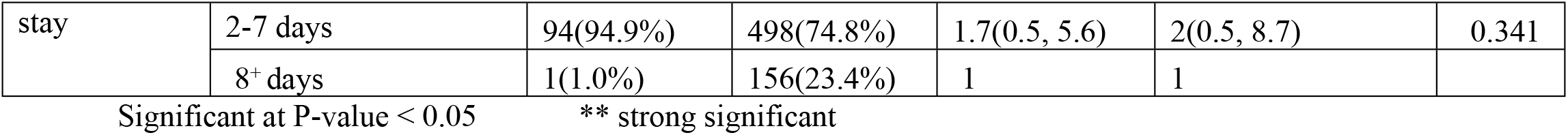
Multivariate logistic regression analysis of maternal and newborn characteristics by early neonatal mortality, NICU of Referral hospital from May 2019-May 2021

### Top leading causes of early neonatal mortality at NICU of Shiek Hassen referral hospital, Somali, Ethiopia

Prenatal Asphyxia was the major cause of early neonatal mortality, followed by prematurity, with 32 (32.3%) and 31 (31.3%) deaths, respectively. Sepsis and congenital anomalies were the third and fourth causes of early neonatal mortality, respectively, 24 (24.2%) and 8 (8.1%) (Table 4).

**Table 4.** Top leading causes of early neonatal mortality at NICU, Referral Hospital from May 2019-May 2021

| Ranks | Causes of early neonatal mortality | Frequency (%) |
| --- | --- | --- |
| 1 | Prenatal Asphyxia | 32(32.3) |
| 2 | Prematurity | 31(31.3) |
| 3 | Sepsis | 24(24.2) |
| 4 | Congenital Malformation | 8(8.1) |
| 5 | Others | 4(4.0) |

## Discussion

The goal of the study was to determine the causes and determinants of early newborn death in a referral hospital in Ethiopia’s Somali region. There were 765 live births admitted throughout the study period, with 99 deaths occurring during the first seven days, resulting in an early neonatal mortality rate of 13 per 1000 live births, or 1.3 percent. The major causes of admission and death in the newborn Intensive Care Unit include birth asphyxia, preterm, sepsis, and congenital abnormalities. This is in line with the findings of other investigations [10, 17, 20, 21].

This study’s early newborn mortality rate of 13 per 1000 live births (1.3%) is similar to a study in Afghanistan that showed 1.4 percent[22] but lower than studies in Northern Ethiopia(1.86%)[23], Bukinafaso [2.6%] [24], Nigeria(3.8%) [25] and shaanxi province, China(7.9%) [26]. The findings are also in line with other researches that show majority of neonatal deaths occur in the first week of life, particularly in the first 24 hours [7-9, 27, 28]. During the neonatal era, the risk of death is highest at the time of birth and gradually diminishes over the following days and weeks. Within the first 24 hours of delivery, up to 36% of neonates die, and approximately 73% die within the first week of life [29].

Prematurity was discovered to be a significant determinants of early newborn mortality. This is in line with the fact that preterm newborns have a greater mortality rate than term newborns[30]. Organ failure, neurodevelopmental and learning disabilities, vision problems, and long-term cardiovascular and non-communicable diseases are all risks for preterm babies [31, 32]. The findings of this study are likewise consistent with those of Ethiopian and Sub-Saharan African studies [7, 8, 10, 18].

Another important determining factor that exhibited a protective effect against early newborn mortality was neonatal resuscitation. This is in line with the findings of a Delphi panel of 18 experts, who indicated that urgent newborn facility-based resuscitation would avoid an additional 10% of preterm deaths, while community-based resuscitation would prevent an additional 20% of intrapartum-related and 5% of preterm fatalities[33]. Several other studies of neonatal resuscitation in low- and middle-income countries have shown that it has the potential to save newborn lives [34-36].

Hypothermia (lower than normal body temperature) has also been linked to early newborn death. This is consistent with researches conducted in Ethiopia and elsewhere, which found that neonatal admission hypothermia dramatically increases the chance of death. [37, 38]. Hypothermia during NICU admission increased the risk of early newborn death, according to another study [39, 40]. This could be linked to procedures such as delivering babies at <25°C(delivery room temperature), providing respiratory assistance with cold air during transfer to the NICU, and not using a cap on newborns, as well as unnecessary delays in skin-to-skin contact, preterm, and significant bacterial infection [40-42]. Furthermore, the majority (442, or 87.7%) of hypothermic neonates admitted to the study hospital did not receive Kangroo mother care (KMC), which protects the newborn from infection, effectively treats hypothermia, improves gastrointestinal function and cardiorespiratory stability, and encourages breast feeding[43] and thus reduces early neonatal mortality [44, 45].

A shorter stay in the NICU (less than 24 hours) was also discovered to be a robust determinant of early newborn mortality—a risk effect. This suggests that neonates who were discharged within 24 hours had a higher chance of mortality than those who stayed in the NICU for longer. The finding is in line with a research in Jimma and eastern Ethiopia which found that shorter durations (less than 3 and 7 days) and longer durations (> 5 days) are risk factors for infant mortality [10, 39, 46]. However, this finding is in conflict with a research conducted in Ethiopia’s Somali region, which found that an average duration of stay of less than 2 days was protective against newborn mortality [8].

The other determinant of early newborn mortality discovered in this study was maternal death; newborns whose mothers died were more likely to die; the early post-natal period is a dangerous time for both mothers and their babies, and is strongly related to labor, intra-partum, and immediate new-born care practices. Studies have found a handful of maternal factors for early newborn death, such as hemorrhage and pregnancy-induced hypertension [7, 47-51]. Furthermore, poor-quality care is responsible for 61% of neonatal deaths and half of maternal mortality[52]. This is because newborn care cannot be offered in isolation; it must be provided in conjunction with high-quality maternal care, which is also critical in saving lives. The health of mothers and their babies is intertwined, and providing appropriate interventions has the potential to save 71 percent of newborn fatalities, 33 percent of stillbirths, and 54 percent of maternal deaths if implemented fully[53].

This study has the advantage of reviewing all newborns admitted to the hospital’s NICU from 2019 to 2021, eliminating any potential sampling error. However, because the study used secondary data, the findings are subjected to concerns such as incomplete recording, which were mitigated during data collecting and analysis. It also doesn’t represent neonatal mortality at community level.

## Conclusions

The magnitude of early neonatal mortality is considerable in Newborn Intensive Care Unit and causes of newborn deaths are preventable. Enhancing quality of care including infection prevention and hypothermia through mentorship and encouraging the KMC practice is of paramount important NICU of the Hospital.

## Data Availability

All relevant data are within the manuscript and its Supporting Information files.

## Supporting information

S1 file. SPSS dataset

## Acknowledgement

We appreciate the assistance of SHYR Hospital officials and NICU nurses throughout data collecting. We also appreciate the help of data collectors, supervisors, and Jigjiga University during data collectecting.

## Author Contributions

**Conceptualization:** Ahmed Tahir Ahmed, Abdifatah Elmi Farah, Hussein Nooh Ali

**Data curation:** Ahmed Tahir Ahmed, Abdifatah Elmi Farah

**Formal analysis:** Ahmed Tahir Ahmed, Abdifatah Elmi Farah

**Methodology:** Ahmed Tahir Ahmed

**Supervision:** Ahmed Tahir Ahmed, Hussein Nooh Ali, Muse Obsiye Ibrahim

**Writing – original draft:** Ahmed Tahir Ahmed, Abdifatah Elmi Farah.

**Writing – review & editing:** Ahmed Tahir Ahmed, Abdifatah Elmi Farah, Hussein Nooh Ali, Muse Obsiye Ibrahim

